# Interactions among common non-SARS-CoV-2 respiratory viruses and influence of the COVID-19 pandemic on their circulation in New York City

**DOI:** 10.1101/2021.10.14.21264861

**Authors:** Haokun Yuan, Alice Yeung, Wan Yang

**Author notes:** Correspondence to: Wan Yang, Department of Epidemiology, Mailman School of Public Health Columbia University, 722 W 168th Street, Room 514, New York, NY 10032.

## Abstract

**Background:** Non-pharmaceutical interventions (NPIs) and voluntary behavioral changes during the COVID-19 pandemic have influenced the circulation of non-SARS-CoV-2 respiratory infections. We aimed to examine interactions among common non-SARS-CoV-2 respiratory virus and further estimate the impact of the COVID-19 pandemic on these viruses.

**Methods:** We analyzed incidence data for seven groups of respiratory viruses in New York City (NYC) during Oct 2015 - May 2021 (i.e., before and during the COVID-19 pandemic). We first used elastic net regression to identify potential virus interactions and further examined the robustness of the found interactions by comparing the performance of Auto Regressive Integrated Moving Average (ARIMA) models with and without the interactions. We then used the models to compute counterfactual estimates of cumulative incidence and estimate the reduction during the COVID-19 pandemic period from March 2020 to May 2021, for each virus.

**Results:** We identified potential interactions for three endemic human coronaviruses (CoV-NL63, CoV-HKU, and CoV-OC43), parainfluenza (PIV)-1, rhinovirus, and respiratory syncytial virus (RSV). We found significant reductions (by ~70-90%) in cumulative incidence of CoV-OC43, CoV-229E, human metapneumovirus, PIV-2, PIV-4, RSV, and influenza virus during the COVID-19 pandemic. In contrast, the circulation of adenovirus and rhinovirus was less affected.

**Conclusions:** Circulation of several respiratory viruses has been low during the COVID-19 pandemic, which may lead to increased population susceptibility. It is thus important to enhance monitoring of these viruses and promptly enact measures to mitigate their health impacts (e.g., influenza vaccination campaign and hospital infection prevention) in the coming months.

## Introduction

Viral respiratory infections are one of the leading causes of disease in humans. Each year, numerous respiratory viruses co-circulate in the population, causing substantial public health burden [1] and economic loss [2, 3]. Previous studies have suggested that respiratory viruses may interfere with and change the risk, timing, or natural history of infection of one another [4]. For instance, in 2009, seasonal epidemic of respiratory syncytial virus (RSV) in Israel was temporarily delayed due to the A(H1N1) pandemic [5]. Potential mechanisms including competitions within hosts (e.g., infecting cells) and population-level interactions have been proposed to explain such virus interactions [4]. However, the specific interactions among different respiratory viruses and the impact on their collective transmission dynamics have not been well characterized.

Before the COVID-19 pandemic, influenza was the foremost public health concern among all respiratory infections. As such, much research effort has been devoted to understand the transmission dynamics of influenza viruses and their interactions with other respiratory viruses [4]. In contrast, milder infections such as human endemic coronaviruses have received far less attention. In addition, previous studies tend to ignore the different subtypes of respiratory viruses and only examine interactions at the level of genus. However, subtypes from the same virus group may have different seasonality (e.g., the four subtypes of parainfluenza viruses) and competitions within genus tend to be more intense [e.g., influenza A(H1N1) and A(H3N2)][6]. As such, combining all subtypes of a virus as a whole regardless of their circulation patterns may mask the true interactions.

Following its emergence in late 2019, SARS-CoV-2 has spread to 214 countries and territories, causing the COVID-19 pandemic [7]. The widespread prevalence of SARS-CoV-2 may affect the circulation of other respiratory viruses, via virus interactions. In addition, during the COVID-19 pandemic, non-pharmaceutical interventions (NPIs) such as social distancing, school closures, travel bans, and mask-wearing that aimed to mitigate COVID- 19 transmission had also limited the transmission of other respiratory viruses. Seasonal respiratory viruses such as influenza were found to be at low circulation during the 2020 respiratory virus season amid the COVID-19 pandemic [8, 9]. However, to what extent the COVID-19 pandemic has impacted the circulation of common respiratory viruses and potential differences by virus remains unclear.

In this work, we analyzed incidence data for seven groups of respiratory viruses in New York City (NYC) before and during the COVID-19 pandemic. We first used elastic net regression to identify potential interactions among the viruses at the subtype level (13 in total). We further hypothesized that strong interactions should allow models incorporating the relationship to more accurately predict the incidence of related respiratory viruses. To examine the robustness of the found interactions, we thus built and compared the performance of Auto Regressive Integrated Moving Average (ARIMA) models with and without the interactions. Lastly, we used the best-performing models to estimate the impact of the COVID-19 pandemic on circulation of each of the 13 respiratory viruses.

## Methods

### Virus surveillance data

The virus surveillance data were collected from a subset of NYC participating laboratories that test for multiple respiratory viruses in addition to influenza and RSV. Tested viruses included adenovirus (Adv), coronavirus (CoV), human metapneumovirus (HMPV), rhinovirus (RV), parainfluenza (PIV), RSV, and influenza virus (IV) overall and by subtype (Table 1). The data included the number of respiratory pathogen panel tests requested each week and the number tested positive for each virus during Week 40 of 2015 to Week 20 of 2021. Prior to the COVID-19 pandemic, the total number of samples tested each respiratory virus season (defined as the 40^th^ week of the year to the 39^th^ week the next year) increased over time with the expansion of testing. To account for this time trend, we adjusted the weekly incidence by multiplying the ratio of the number of samples tested during the season to that number in season 2015-2016 (the first season of data collection). For the COVID-19 pandemic period (week 10 of 2020 to week 20 of 2021; note the first COVID-19 case was reported in NYC during week 10 of 2020 [10]), the number of samples tested each week fluctuated substantially from week to week. To account for this short-term fluctuation, we adjusted the incidence during COVID-19 pandemic period week by week relative to the corresponding week during season 2015- 2016.

**Table 1.**
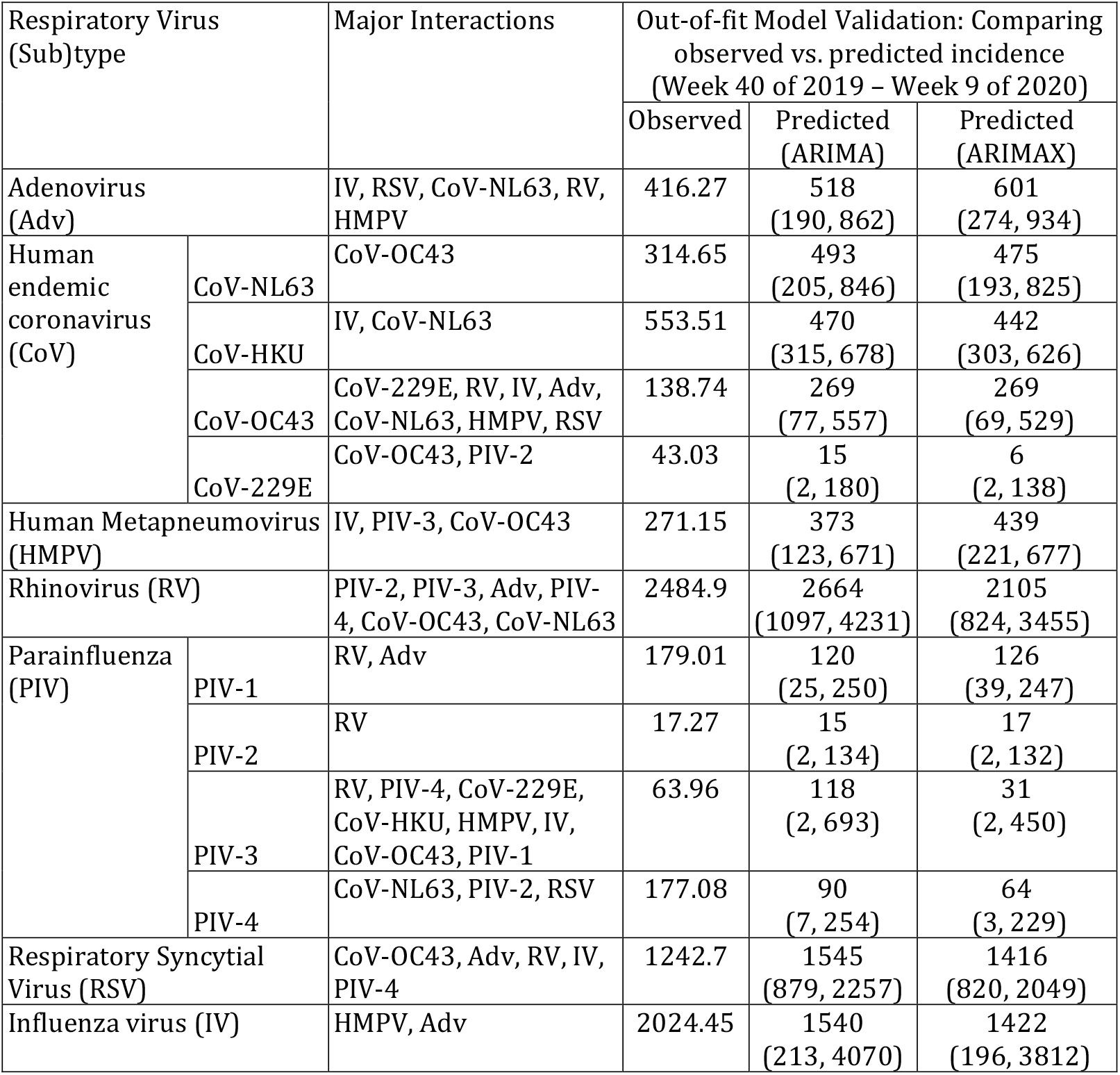
Model-identified potential viral interactions during the pre-COVID-19 period. Column “Major Interactions” show identified interactions from the initial selection by the elastic net regression and the stepwise forward selection for the ARIMAX model (i.e., these were the final variables included in the ARIMAX model). Estimated strengths of interactions are shown in Table S1. The 3^rd^ panel shows comparison between the observed and model-predicted cumulative incidence (mean and prediction intervals in parentheses) during the testing period (i.e., Week 40 of 2019 – Week 9 of 2020); the prediction intervals should include the observed if the model is accurate, which is the case for all models here.

| Respiratory Virus (Sub)type |  | Major Interactions | Out-of-fit Model Validation: Comparing observed vs. predicted incidence (Week 40 of 2019 – Week 9 of 2020) |  |  |
| --- | --- | --- | --- | --- | --- |
|  |  |  | Observed | Predicted (ARIMA) | Predicted (ARIMAX) |
| Adenovirus (Adv) |  | IV, RSV, CoV-NL63, RV, HMPV | 416.27 | 518<br>(190, 862) | 601<br>(274, 934) |
| Human endemic coronavirus (CoV) | CoV-NL63 | CoV-OC43 | 314.65 | 493<br>(205, 846) | 475<br>(193, 825) |
|  | CoV-HKU | IV, CoV-NL63 | 553.51 | 470<br>(315, 678) | 442<br>(303, 626) |
|  | CoV-OC43 | CoV-229E, RV, IV, Adv, CoV-NL63, HMPV, RSV | 138.74 | 269<br>(77, 557) | 269<br>(69, 529) |
|  | CoV-229E | CoV-OC43, PIV-2 | 43.03 | 15<br>(2, 180) | 6<br>(2, 138) |
| Human Metapneumovirus (HMPV) |  | IV, PIV-3, CoV-OC43 | 271.15 | 373<br>(123, 671) | 439<br>(221, 677) |
| Rhinovirus (RV) |  | PIV-2, PIV-3, Adv, PIV-4, CoV-OC43, CoV-NL63 | 2484.9 | 2664<br>(1097, 4231) | 2105<br>(824, 3455) |
| Parainfluenza (PIV) | PIV-1 | RV, Adv | 179.01 | 120<br>(25, 250) | 126<br>(39, 247) |
|  | PIV-2 | RV | 17.27 | 15<br>(2, 134) | 17<br>(2, 132) |
|  | PIV-3 | RV, PIV-4, CoV-229E, CoV-HKU, HMPV, IV, CoV-OC43, PIV-1 | 63.96 | 118<br>(2, 693) | 31<br>(2, 450) |
|  | PIV-4 | CoV-NL63, PIV-2, RSV | 177.08 | 90<br>(7, 254) | 64<br>(3, 229) |
| Respiratory Syncytial Virus (RSV) |  | CoV-OC43, Adv, RV, IV, PIV-4 | 1242.7 | 1545<br>(879, 2257) | 1416<br>(820, 2049) |
| Influenza virus (IV) |  | HMPV, Adv | 2024.45 | 1540<br>(213, 4070) | 1422<br>(196, 3812) |

This study was classified as public health surveillance and exempt from ethical review and informed consent by the Institutional Review Boards of both Columbia University and NYC DOHMH.

### Selection of key viral interactions during the pre-COVID period

We used elastic net regression models [11] to identify, for each virus, the set of other viruses consistently identified as interacting viruses, during Week 40 of 2015 – Week 9 of 2020, i.e. before the first COVID-19 case was reported in NYC [10]. To avoid spurious correlation due to a common winter-time seasonality shared by some viruses [12], we first used a linear regression model to identify and remove the seasonal trend for each virus. The model took the following form:

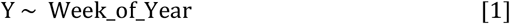

where Y is the adjusted weekly incidence (see the Data section) and Week_of_Year is an indicator variable for each week of the calendar year (1:52 for annual seasonal cycle and 1:104 for biennial cycles). For each virus, we fitted both annual and biennial cycles and used the adjusted R^2^ to determine the most likely seasonal cycle for each virus for removal of seasonal trend.

We then used the detrended time series (i.e., the residuals after removing the seasonal trend) for the elastic net regression model. Similar to lasso (i.e., least absolute shrinkage and selection operator) and ridge regressions, elastic net shrinks regression coefficients by imposing a penalty on their size. However, instead of penalizing by the sum of absolute coefficients (L1—lasso penalty) or the sum-of-squares coefficients (L2—ridge penalty), elastic net penalizes with both L1 and L2 and is formulated as:

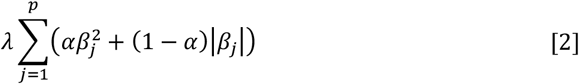

where λ controls the amount of shrinkage, α controls the distribution between L1 and L2, and *β*_*j*_’s represent the regression coefficients that minimize the penalty (i.e. formula 2). Since elastic net shares traits of both ridge and lasso regression, while it selects variables like lasso, it also allows coefficients of correlated predictors to shrink together and provides a more stable selection result.

We fitted 500 elastic net regressions with 10-fold cross-validation and pooled all interactions selected at least in half of the 500 runs for further testing (see next section). Thirteen models were developed, one model each for Adv, HMPV, RV, RSV, and IV, and for each of four subtypes of CoV and PIV. For influenza, due to the more erratic circulation pattern of different types and subtypes and short study period with available data (i.e., five years), we combined all types and subtypes.

### Testing the identified interactions

To further test the identified interactions from the elastic net regression models, we examined if inclusion of any of the found interactions in an ARIMA model with exogenous variables (ARIMAX) model [13] improves model fit. Specifically, we used forward stepwise selection to eliminate exogenous variables (i.e. independent variables; here the interactions identified by the elastic net regression) that do not improve model fit, based on the Bayesian information criterion (BIC) [14]. That is, the ARIMAX model with the lowest BIC is deemed as the best-fit model. For comparison, we also fit the data (without the time series for the identified interacting viruses) to an ARIMA model. Model with the lower relative root mean squared error (rRMSE) was deemed to have a better fit. The ARIMA model took the following form:

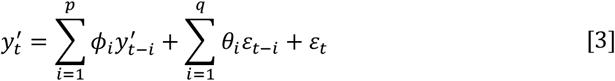

where *y*^′^ is a differenced time series with differencing degree *d*; *p* is the order of autoregressive (AR) model; *q* is the order of moving average (MA) model; *ϕ*_*i*_ (*i* = 1, …, *p*) are the coefficients for the AR terms; *θ*_*i*_ (*i* = 1, …*q*) are the coefficients for the MA terms; and ε is the error term.

Similarly, the ARIMAX model was formulated as:

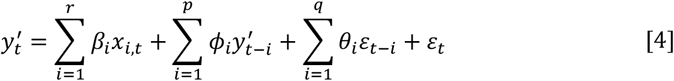

where *x*_*i*_ (*i* = 1, …, *r*) are the exogenous variables and *β*_*i*_ (*i* = 1, …, *r*) the corresponding coefficients.

### Model validation

We tested the ARIMA and ARIMAX models using data prior to the COVID-19 pandemic (i.e. Week 40 of 2015 – Week 9 of 2020). Specifically, we divided this pre-COVID-19 dataset into a training (Week 40 of 2015 – Week 39 of 2019, i.e. 4 full respiratory seasons) and a testing (Week 40 of 2019 – Week 9 of 2020, i.e. the last respiratory season before the COVID-19 pandemic) subset. The models (either ARIMA or ARIMAX) were first fit to the training subset; the trained models were then used to predict the weekly incidence for each virus during the testing period for out-of-fit model validation. We compared model performance based on rRMSE during the testing period. In addition, as the weekly incidence tended to be low, we also evaluated the models based on the cumulative incidence during the testing period and the 95% prediction interval; if the observed cumulative incidence fell within the 95% prediction interval, the model was deemed accurate.

### Estimating the impact of COVID-19 pandemic on circulation of non-SARS-CoV-2 viruses

We used the validated models (ARIMA or ARMIAX) to generate counterfactual estimates for each virus during the pandemic period – i.e., the expected cumulative incidence should there be no pandemic. To enhance model performance, we refitted the validated models using data during the entire pre-COVID-19 period (i.e., through Week 9 of 2020) and used them to predict the incidence for each virus during the COVID-19 period (here Week 10 of 2020 – Week 20 of 2021). We then compared the model counterfactual estimates of cumulative incidence during the COVID-19 period (*C*_*counterfactual*_) to the observations (*C*_*observed*_) to estimate the impact of the COVID-19 pandemic. We computed the percent reduction in cumulative incidence due to the COVID-19 pandemic for each virus as:

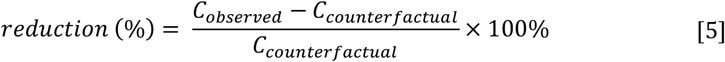

## Results

### Respiratory virus circulation before and after the introduction of SARS-CoV-2

During the pre-COVID-19 period, influenza viruses were the most commonly detected in our dataset (up to 400 adjusted case counts per week), followed by RV (up to ~250 per week), CoV and RSV (both up to ~150 per week); in comparison, other viruses (HMPV, Adv, and PIV) tended to have low cases detected (around 40 – 60 cases during peak weeks; Fig S1). Most of the respiratory viruses (IV, Adv, CoV, RSV, and HMPV) included here had outbreaks in the winter every year (Figs 1–2). In contrast, RV cases were detected year-round and tended to have two comparable seasonal outbreaks each year — one in the winter and one in the summer (Fig 1). PIV cases were also detected throughout the year, but the outbreak patterns were less obvious (Fig 3). For the same virus group, different subtypes tended to alternate in circulation and recurred biennially with irregular peaks (e.g., the four coronaviruses in Fig 2; PIV-1 and PIV-2 in Fig 3). In addition, among the four coronaviruses, weekly case counts were highly correlated for virus pairs belonging to different genera (*r* = 0.82 between CoV-OC43 and CoV-229E and 0.76 between CoV-NL63 and CoV-HKU). In contrast, the circulation of different influenza types and subtypes and PIV-4 appeared less regular, with few cases detected in some years.

**Fig. 1.**
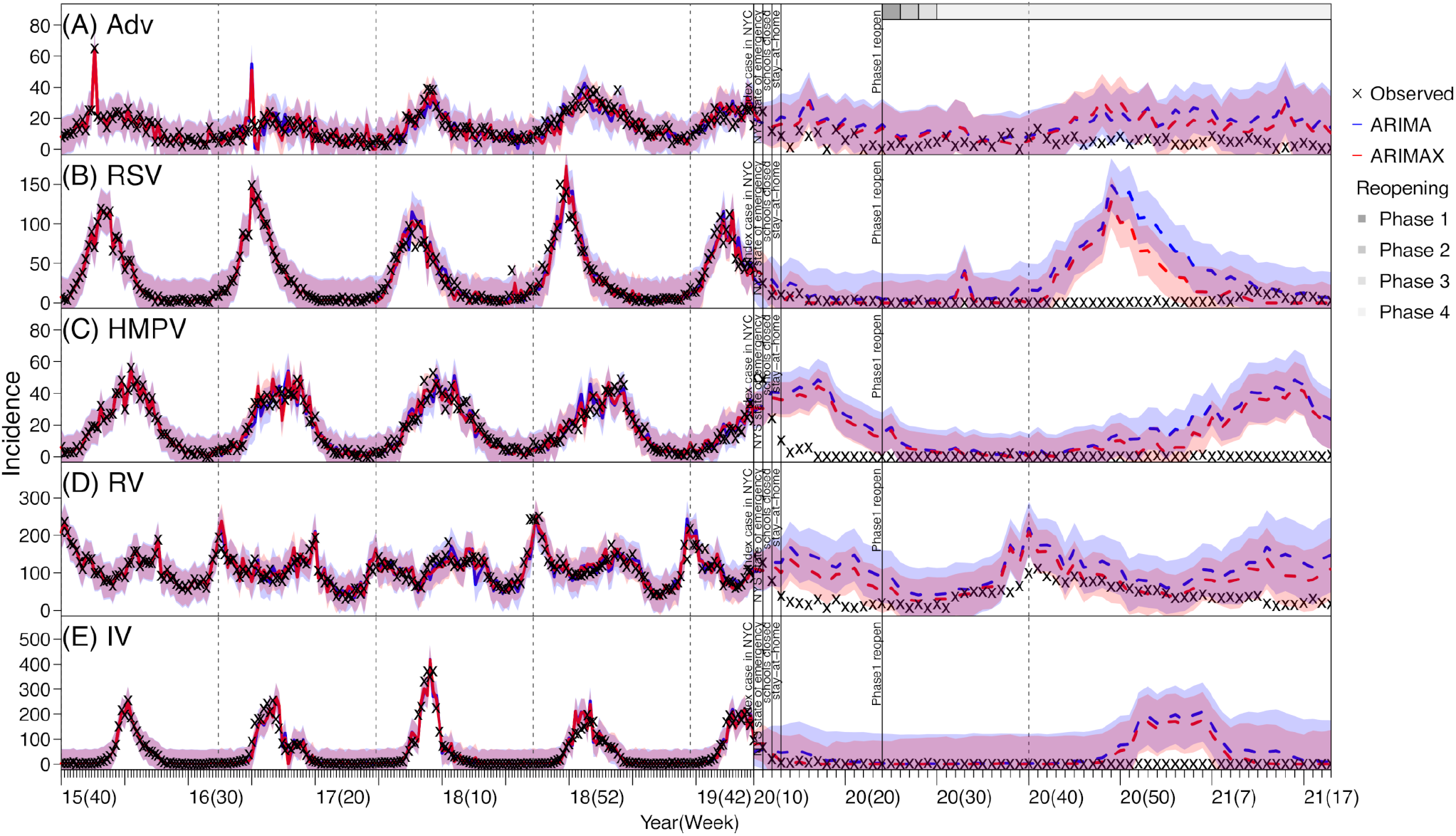
Model fit and prediction of weekly incidence for adenovirus (Adv; A), respiratory syncytial virus (RSV; B), human metapneumovirus (HMPV; C), rhinovirus (RV; D), and influenza virus (IV; E) using ARIMA and ARIMAX models. Crosses (‘x’) show scaled weekly incidence. Blue lines (mean) and shaded areas (95% confidence intervals) show model fit (solid lines) and prediction (dashed lines) using the ARIMA models; red lines (mean) and shaded areas (95% confidence intervals) show model fit (solid lines) and prediction (dashed lines) using the ARIMAX models. Vertical dashed lines indicate the start of each respiratory virus season and vertical black lines mark timing of major COVID-19 events; grey bars on the top of the plot indicate different reopening phases in NYC. See criteria of reopening and phases at https://forward.ny.gov

**Fig. 2.**
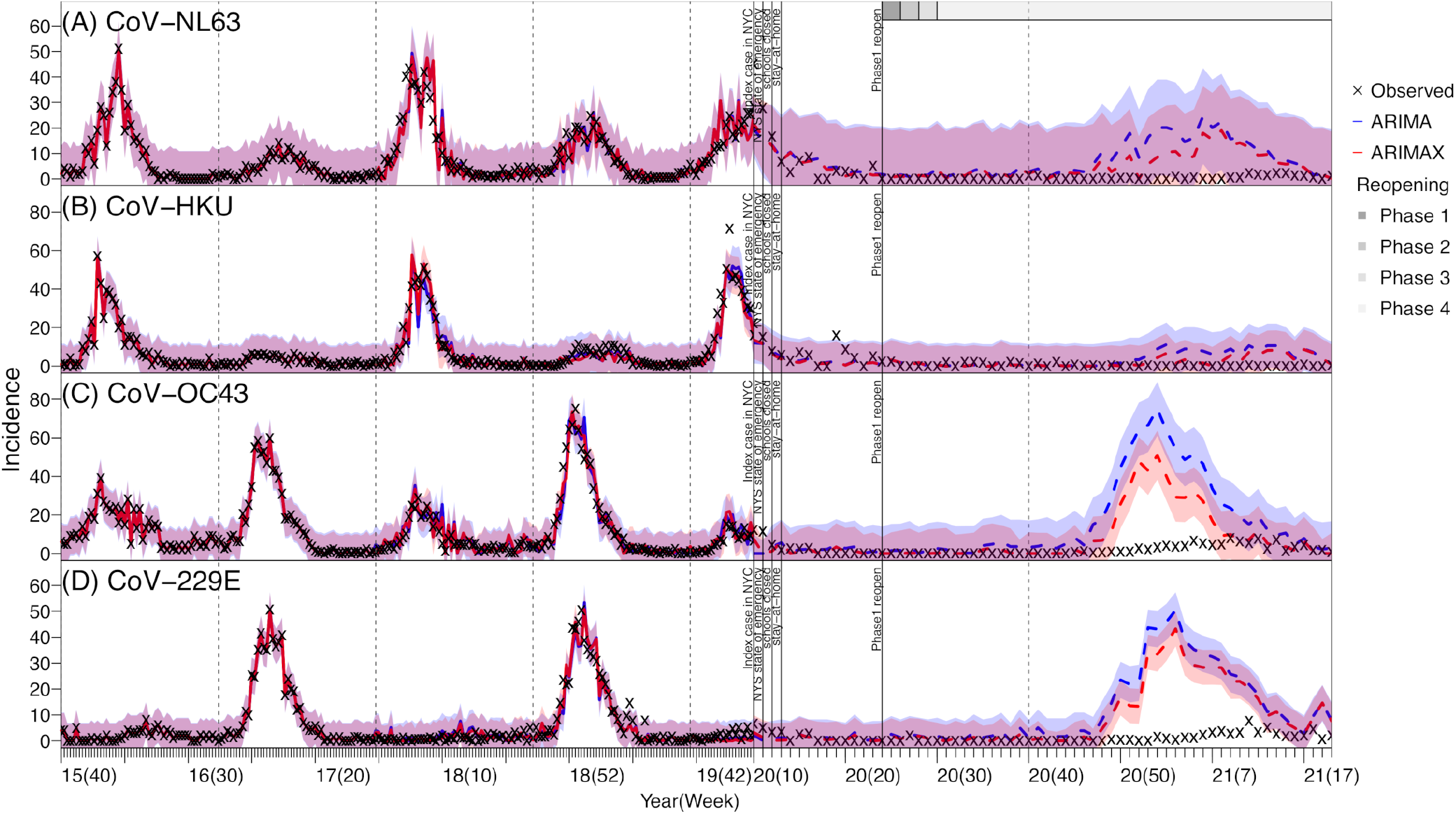
Model fit and prediction of weekly incidence for human endemic coronaviruses: Cov-NL63 (A), Cov-HKU (B), Cov-OC43 (C), and Cov-229E (D) using ARIMA and ARIMAX models. Blue lines (mean) and shaded areas (95% confidence intervals) show model fit (solid lines) and prediction (dashed lines) using the ARIMA models; red lines (mean) and shaded areas (95% confidence intervals) show model fit (solid lines) and prediction (dashed lines) using the ARIMAX models. Vertical dashed lines indicate the start of each respiratory virus season and vertical black lines mark timing of major COVID-19 events; grey bars on the top of the plot indicate different reopening phases in NYC. See criteria of reopening and phases at https://forward.ny.gov

**Fig. 3.**
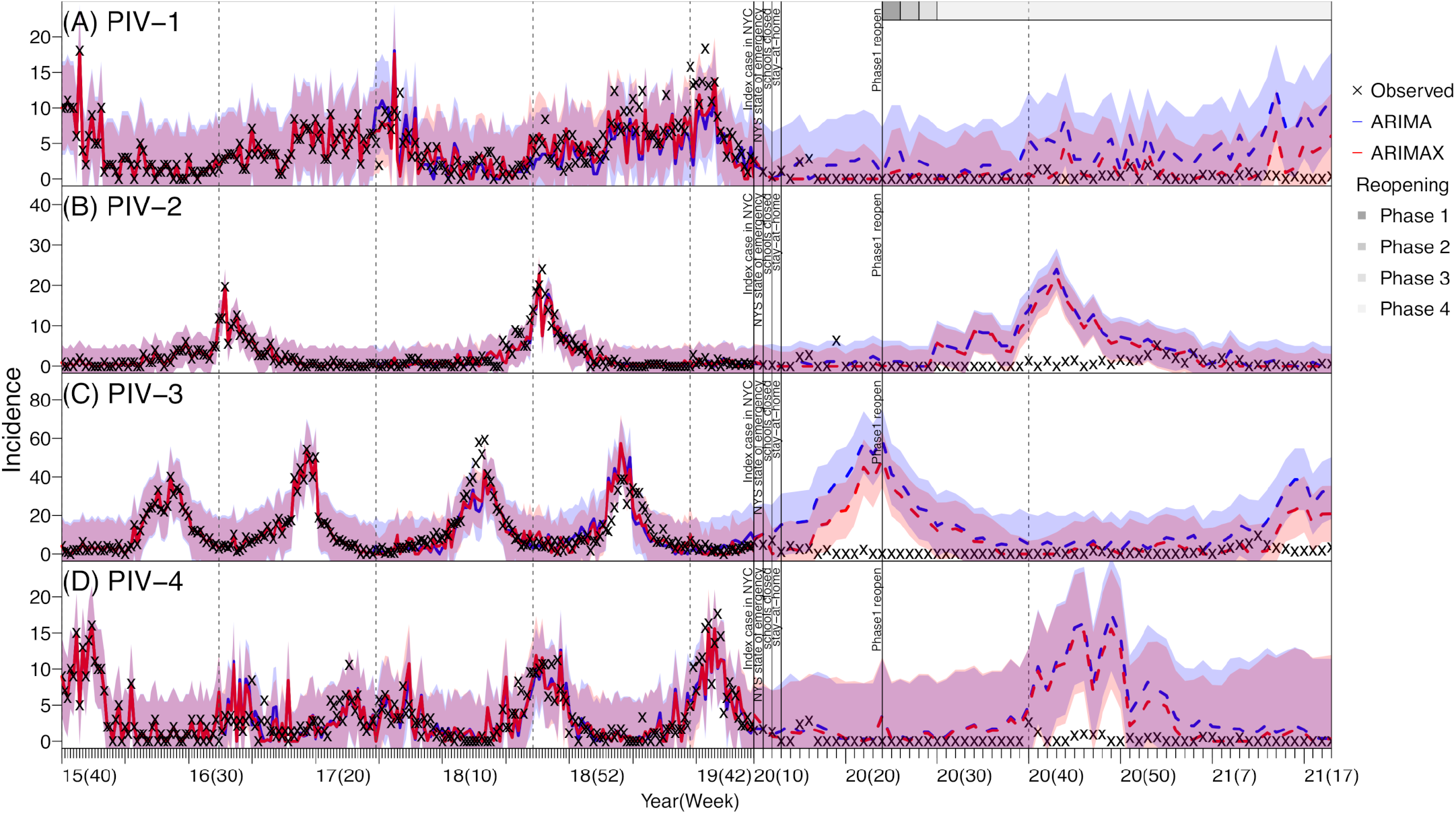
Model fit and prediction of weekly incidence for parainfluenza viruses: PIV-1 (A), PIV-2 (B), PIV-3 (C), and PIV-4 (D) using ARIMA and ARIMAX models. Blue lines (mean) and shaded areas (95% confidence intervals) show model fit (solid lines) and prediction (dashed lines) using the ARIMA models; red lines (mean) and shaded areas (95% confidence intervals) show model fit (solid lines) and prediction (dashed lines) using the ARIMAX models. Vertical dashed lines indicate the start of each respiratory virus season and vertical black lines mark timing of major COVID-19 events; grey bars on the top of the plot indicate different reopening phases in NYC. See criteria of reopening and phases at https://forward.ny.gov

### Potential virus interactions and the impact on model predictability

The elastic net regression models identified several potential associations among the respiratory viruses (Table 1 and Table S1). However, patterns of found interactions were not readily clear and estimated strengths were in general low (all coefficients ≤0.2; Table S1). Further examination using the time series models indicated that, for three coronaviruses (i.e., CoV-NL63, CoV-HKU, and CoV-OC43), RV, PIV-1, and RSV, the inclusion of the identified interacting viruses in the ARIMAX model generated more accurate predictions than the ARIMA model (Table S2). However, this improvement was not substantial (1.6% – 15.4% reduction in rRMSE, see Table S2), likely because epidemics of these respiratory viruses were strongly driven by their underlying seasonality. For the remaining viruses, the inclusion of the interactions did not improve the performance of the ARIMAX model. Overall, both time series models were able to accurately predict the epidemic trajectories (Figs S2–4) as well as the cumulative incidence during the testing period (Week 40 of 2019 – Week 9 of 2020, i.e., the main part of the last respiratory virus season before the COVID-19 pandemic; Table 1). As such, below we present results from both models.

### Impact of the COVID-19 pandemic on non-SARS-CoV-2 viruses

The majority of the respiratory viruses included here appeared to have had lower circulation during the COVID-19 pandemic than would be expected (comparing the model predicted and the observed incidence in Figs 1–3). This is likely due to the continued NPIs implemented in NYC during March 2020 – May 2021 (the end of our study period; see Figs 1–3 for major NPIs implemented). In particular, the circulation of CoV-OC43, CoV-229E, HMPV, PIV-2, PIV-4, RSV, and IV all reduced significantly during the COVID-19 pandemic period — the observed cumulative incidence during this period fell outside the model predicted 95% intervals and the estimated mean reduction was around 70-90% for these viruses (Table 2). In contrast, Adv and RV appeared to be less affected (Fig 1 A and D). Incidence of these two viruses, while low, was nonzero during most weeks of the pandemic period (59 and 63 out of 63 weeks, respectively); and for RV, incidence increased substantially during the summer of 2020, despite the implemented NPIs (Fig 2D). For CoV-NL63 and CoV-HKU, the models did not detect a significant reduction, likely because epidemics of these viruses tended to recur every other year and their lull periods coincided with the pandemic period (Fig. 2 A and B).

**Table 2.** Estimated impact of the COVID-19 pandemic on circulation of non-SARS-CoV-2 viruses in NYC. Column “Observed” shows the scaled cumulative incidence as recorded during the COVID-19 period (March 2020 – May 2021) for each virus (listed in column “Respiratory Virus (Sub)type”). The “Predicted” columns show model-predicted mean (and 95% prediction intervals) using the ARIMA or ARIMAX model (specified on the row above) and the “% Reduction” columns show estimated percent reduction (mean and 95% confidence interval) during the COVID-19 pandemic period, per Eqn 5. Significant reductions are bolded.

| Respiratory Virus<br>(Sub)type |  | Cumulative Incidence during the COVID-19 period |  |  |  |  |
| --- | --- | --- | --- | --- | --- | --- |
|  |  | Observed | ARIMA model<br>counterfactual estimates |  | ARIMAX model<br>counterfactual estimates |  |
|  |  |  | Predicted | % Reduction | Predicted | % Reduction |
| Adenovirus<br>(Adv) |  | 360.33 | 1061<br>(146, 2185) | -66%<br>(-84%, 147%) | 930<br>(114, 2056) | -61%<br>(-82%, 216%) |
| Human<br>endemic<br>coronavirus<br>(CoV) | CoV-NL63 | 148.49 | 495<br>(25, 1679) | -70%<br>(-91%, 494%) | 363<br>(16, 1516) | -59%<br>(-90%, 828%) |
|  | CoV-HKU | 124.09 | 246<br>(4, 975) | -50%<br>(-87%, 3002%) | 165<br>(7, 820) | -25%<br>(-85%, 1673%) |
|  | CoV-OC43 | 132.53 | 921<br>(505, 1790) | **-86%**<br>**(-93%, -74%)** | 504<br>(239, 1247) | **-74%**<br>**(-89%, -45%)** |
|  | CoV-229E | 78.36 | 675<br>(431, 1125) | **-88%**<br>**(-93%, -82%)** | 535<br>(336, 950) | **-85%**<br>**(-92%, -77%)** |
| Human<br>Metapneumovirus<br>(HMPV) |  | 154.72 | 1278<br>(591, 2197) | **-88%**<br>**(-93%, -74%)** | 1051<br>(512, 1829) | **-85%**<br>**(-92%, -70%)** |
| Rhinovirus (RV) |  | 2709.14 | 7045<br>(2472, 11837) | -62%<br>(-77%, 10%) | 5414<br>(1754, 9398) | -50%<br>(-71%, 54%) |
| Parainfluenza<br>(PIV) | PIV-1 | 23.51 | 227<br>(23, 647) | -90%<br>(-96%, 2%) | 68<br>(3, 396) | -65%<br>(-94%, 684%) |
|  | PIV-2 | 52.05 | 290<br>(122, 614) | **-82%**<br>**(-92%, -57%)** | 242<br>(105, 534) | **-78%**<br>**(-90%, -50%)** |
|  | PIV-3 | 85.25 | 862<br>(78, 2558) | -90%<br>(-97%, 9%) | 232<br>(2, 1418) | -63%<br>(-94%, 4162%) |
|  | PIV-4 | 20.43 | 237<br>(50, 762) | **-91%**<br>**(-97%, -59%)** | 189<br>(37, 735) | **-89%**<br>**(-97%, -45%)** |
| Respiratory Syncytial<br>Virus (RSV) |  | 250.3 | 1814<br>(561, 4205) | **-86%**<br>**(-94%, -55%)** | 1453<br>(519, 3507) | **-83%**<br>**(-93%, -52%)** |
| Influenza (IV) |  | 258.09 | 2924<br>(584, 10831) | **-91%**<br>**(-98%, -56%)** | 1941<br>(424, 9033) | **-87%**<br>**(-97%, -39%)** |

## Discussion

In this study, we utilized viral surveillance data collected in NYC before and during the COVID-19 pandemic to examine potential interactions among seven groups of respiratory viruses and the impact of the COVID-19 pandemic on their circulations in the population. We identified several potential interactions, particularly, for three coronaviruses (CoV-NL63, CoV-HKU, CoV-OC43), PIV-1, RV, and RSV. In addition, we found a significantly lower number of cases were detected for several viruses (i.e., CoV-OC43, CoV-229E, HMPV, PIV-2, PIV-4, RSV, and IV), suggesting reduced circulation of these viruses, during the COVID-19 pandemic.

While most respiratory viruses had reduced circulation in NYC during the COVID-19 pandemic, epidemic dynamics of individual viruses differed. Influenza viruses, human endemic coronaviruses, and RSV typically circulate during cold months (from late fall to the early spring the next year in NYC). Thus, circulations of all three groups of viruses were in decline at the start of the first pandemic wave (i.e., March 2020). As a result, the full impact of the COVID-19 pandemic on these viruses did not manifest until the following respiratory virus season (beginning in Oct 2020), during which significant case reductions were found. Influenza cases remained low while RSV began to surface in the spring of 2021 (Fig 1B). In addition to reduced contact among individuals due to social distancing and infection reduction due to mask-wearing, the reduced importation of new virus strains from travelers may have also played a role in the observed outcomes for these viruses. In particular, for influenza, seeding of new A(H3N2) strains emerged globally has been shown to play an important role in starting new epidemics in North America [15]. As such, given the likely high population susceptibility to these viruses after the skipped season, it is important to monitor the circulation of these viruses (particularly, influenza) in the population, severity of cases, concentration in geographical area and/or settings (e.g. congregate facilitates) and age groups as global travel gradually resumes in the coming months. This would enable the implementation of more timely infection mitigation measures (e.g., targeted messaging including vaccination promotion).

For PIV, HMPV, and RV, because their epidemic timing coincided with the first COVID-19 pandemic wave in NYC (spring 2020), all three viruses saw the most dramatic case declines in the first few months of the pandemic. However, as NYC partially reopened in the summer of 2020, the epidemic trajectories of these viruses evolved differently, likely due to the differences in infection demographics. Both PIV and HMPV predominantly infect young children [16, 17] and, to a lesser extent, older adults [18, 19]. As such, their transmission was reduced to minimal levels in both key infection age groups when schools closed and intergenerational interactions (i.e., between grand-children and grandparents) reduced during the pandemic. Interestingly, circulation of both viruses remained low after daycares and schools partially reopened, likely due to required preventive measures such as physical distancing and mask-wearing in schools. This low transmission in young children may have in part reduced the risk of transmission to and among older adults. Future work using more detailed data (e.g., household data) may further examine the importance of intergenerational transmission of these viruses and provide insights into infection prevention for older adults, for whom infections could lead to severe health outcomes [18].

For RV, even though viral activities were also lower during the COVID-19 period, case increases were observed during the summer of 2020 when NPIs were relaxed (Fig 1D). Similarly, continued transmission of RV was reported during and after the 2009 influenza A(H1N1) pandemic [20]. Rhinovirus infections occur in most age groups and infection of one serotype confers little immune protection against others [21–23]. This wider infection demographics and the large breadth of RV serotypes (around 160 discovered by 2018 [22]) likely facilitated its transmission locally in the population.

Our study has several limitations. First, the data analyzed here are a subset of all tests done in NYC (i.e., only those from laboratories using the expanded respiratory panel tests) and thus may not be fully representative of the entire population. Second, even though the selection criteria have not changed during the study period, underlying patient characteristics may differ among specimens tested before and during the COVID-19 period, due to changing medical seeking behaviors in response to COVID-19 (e.g., people with mild respiratory symptoms may be more likely to seek testing at the early stage of the pandemic due to concern of COVID-19); this in turn may temporally change the composition of underlying sample population. Third, fewer specimens were tested each week during the early phase of the pandemic due to limited testing supplies and human resources; this reduced sampling likely increased model uncertainty. Fourth, the identified associations (Table 1) were based on population-level epidemic time series and do not imply any causal interactions between each included virus pairs. Future research at the individual level (e.g. frequency of co-infection or subsequent infections by multiple viruses in the same individuals) is warranted to further examine the potential viral interactions reported here. Finally, although we found substantial case reductions during the pandemic for several non-SARS-CoV-2 respiratory viruses, it is difficult to distinguish the impact due to the introduction and circulation of the SARS-CoV-2 virus in the population and that due to the NPIs. Long-term viral surveillance post-pandemic may allow further study on the interactions between SARS-CoV-2 and other respiratory viruses without the presence of NPIs and in turn better understanding of the impact of NPIs on each virus.

## Data Availability

Data are used with permission from the NYC Department of Health and Mental Hygiene under a data use and non-disclosure agreement.

## Acknowledgments

This study was in part supported by the National Institute of Allergy and Infectious Diseases (AI145883). We thank NYC DOHMH colleagues Sharon K. Greene for facilitating the data use agreement for data used in this study and for providing input on the manuscript, Beth Nivin for data collection, Maryam Iqbal for extracting and preparing the viral surveillance data used in this study, and Scott Harper for reviewing the manuscript. We also like to thank all laboratories sharing the data including the following individuals and their institutions: Kenneth Inglima, New York University Langone Medical Center, Shek Man Tsui, Memorial Sloan Kettering Cancer Center, Stephanie D. Vaca, NewYork-Presbyterian/Weill Cornell Medical Center.

## Supplementary Table Captions

**Table S1.** Estimated strengths of virus interactions. For each virus, interacting viruses were identified first by the elastic net regression and further by forward stepwise selection into the final ARIMAX model. Each column shows results for the final ARIMAX model for each of the thirteen viruses included here: non-empty cells indicate viruses in the corresponding rows (see row names) were included and numbers show estimated coefficients in the ARIMAX model (those <0.01 are shown as “0.00” due to rounding) and darker colors indicate stronger interactions; empty cells indicate the corresponding viruses were not included in the ARIMAX model as interacting viruses. For instance, for adenovirus (Adv), five viruses (CoV-NL63, HMPV, RV, RSV and IV) were identified as its interacting viruses; the estimated strengths were highest for HPMV (0.13), followed by RV (0.10) and CoV-NL63 (0.06), and <0.01 for RSV and IV.

|  | Adenovirus | Coronavirus |  |  |  | Human Metapneumovirus | Rhinovirus | Parainfluenza |  |  |  | Respiratory Syncytial Virus | Influenza |
| --- | --- | --- | --- | --- | --- | --- | --- | --- | --- | --- | --- | --- | --- |
|  | Adv | CoV-NL63 | CoV-HKU | CoV-OC43 | CoV-229E | HMPV | RV | PIV-1 | PIV-2 | PIV-3 | PIV-4 | RSV | IV |
| Adv |  |  |  | 0.08 |  |  | 0.05 | 0.11 |  | 0.02 |  | 0.00 | 0.15 |
| CoV-NL63 | 0.06 |  | 0.08 | 0.03 |  |  | 0.12 |  |  |  | 0.00 |  |  |
| CoV-HKU |  |  |  |  |  |  |  |  |  | 0.03 |  |  |  |
| CoV-OC43 |  | 0.09 |  |  | 0.00 | 0.06 | 0.04 |  |  |  |  | 0.00 |  |
| CoV-229E |  |  |  | 0.02 |  |  |  |  |  |  |  |  |  |
| HMPV | 0.13 |  |  | 0.06 |  |  |  |  |  | 0.01 |  | 0.07 | 0.00 |
| RV | 0.10 |  |  | 0.12 |  |  |  | 0.00 | 0.07 | 0.00 |  | 0.11 |  |
| PIV-1 |  |  |  |  |  |  |  |  |  |  |  |  |  |
| PIV-2 |  |  |  |  | 0.14 |  | 0.00 |  |  |  | 0.04 |  |  |
| PIV-3 |  |  |  |  |  | 0.01 | 0.00 |  |  |  |  |  |  |
| PIV-4 |  |  |  |  |  |  | 0.20 |  |  | 0.01 |  |  |  |
| RSV | 0.00 |  |  | 0.04 |  |  |  |  |  |  | 0.04 |  |  |
| IV | 0.00 |  | 0.00 | 0.14 |  | 0.00 |  |  |  |  |  |  |  |

**Table S2.** Model performance during pre-COVID-19 testing period (Oct 2019-Feb 2020). Main interactions were the exogenous variables included in the ARIMAX model. Model performance was measured by relative RMSE and the difference in relative RMSE between the two models is shown in the parentheses.

|  |  | Main Interactions | Relative RMSE<br>(Testing period) |  |
| --- | --- | --- | --- | --- |
|  |  |  | ARIMA | ARIMAX |
| Adenovirus (Adv) |  | IV, RSV, CoV-NL63, RV, HMPV | 0.67 | 0.85<br>(26.87%) |
| Human endemic coronavirus (CoV) | CoV-NL63 | CoV-OC43 | 1.79 | 1.69<br>(-5.59%) |
|  | CoV-HKU | IV, CoV-NL63 | 1.23 | 1.21<br>(-1.63%) |
|  | CoV-OC43 | CoV-229E, RV, IV, CoV-NL63, HMPV, RSV, Adv | 0.73 | 0.66<br>(-9.59%) |
|  | CoV-229E | CoV-OC43, PIV-2 | 0.38 | 0.64<br>(68.42%) |
| Human Metapneumovirus (HMPV) |  | IV, PIV-3, CoV-OC43 | 0.36 | 0.57<br>(58.33%) |
| Rhinovirus (RV) |  | PIV-2, PIV-3, Adv, PIV-4, CoV-OC43, CoV-NL63 | 0.26 | 0.23<br>(-11.54%) |
| Parainfluenza (PIV) | PIV-1 | RV, Adv | 1.19 | 1.02<br>(-14.29%) |
|  | PIV-2 | RV | 0.46 | 0.47<br>(2.17%) |
|  | PIV-3 | RV, PIV-4, CoV-229E, CoV-HKU, HMPV, IV, CoV-OC43, PIV-1 | 0.34 | 0.72<br>(111.76%) |
|  | PIV-4 | CoV-NL63, PIV-2, RSV | 1.8 | 2.24<br>(24.44%) |
| Respiratory Syncytial Virus (RSV) |  | CoV-OC43, Adv, RV, IV, PIV-4 | 0.78 | 0.66<br>(-15.38%) |
| Influenza (IV) |  | HMPV, Adv | 0.85 | 0.96<br>(12.94%) |

## Supplementary Figure Captions

**Fig S1.**
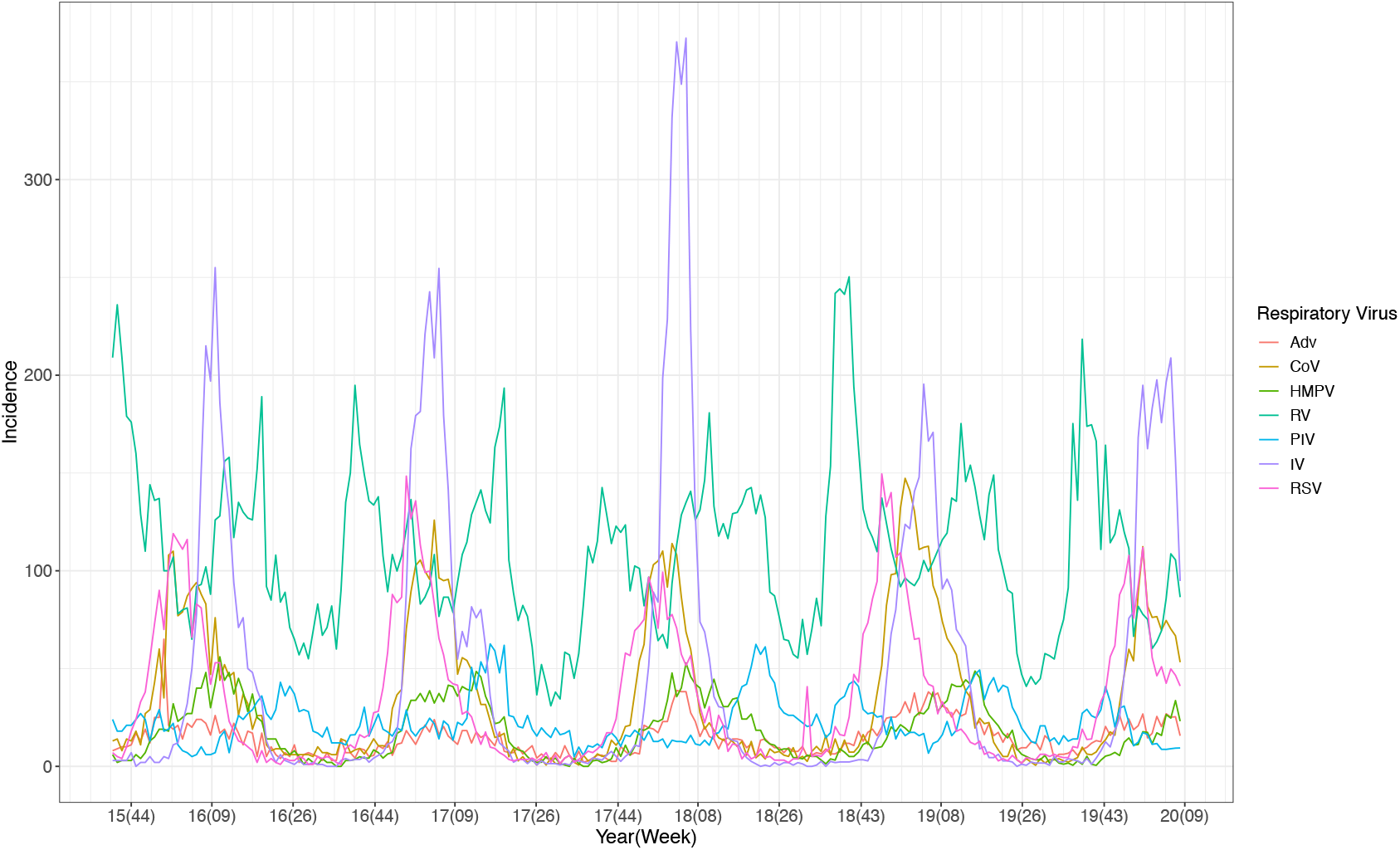
Weekly respiratory virus activities in NYC before COVID-19 pandemic.

**Fig S2.**
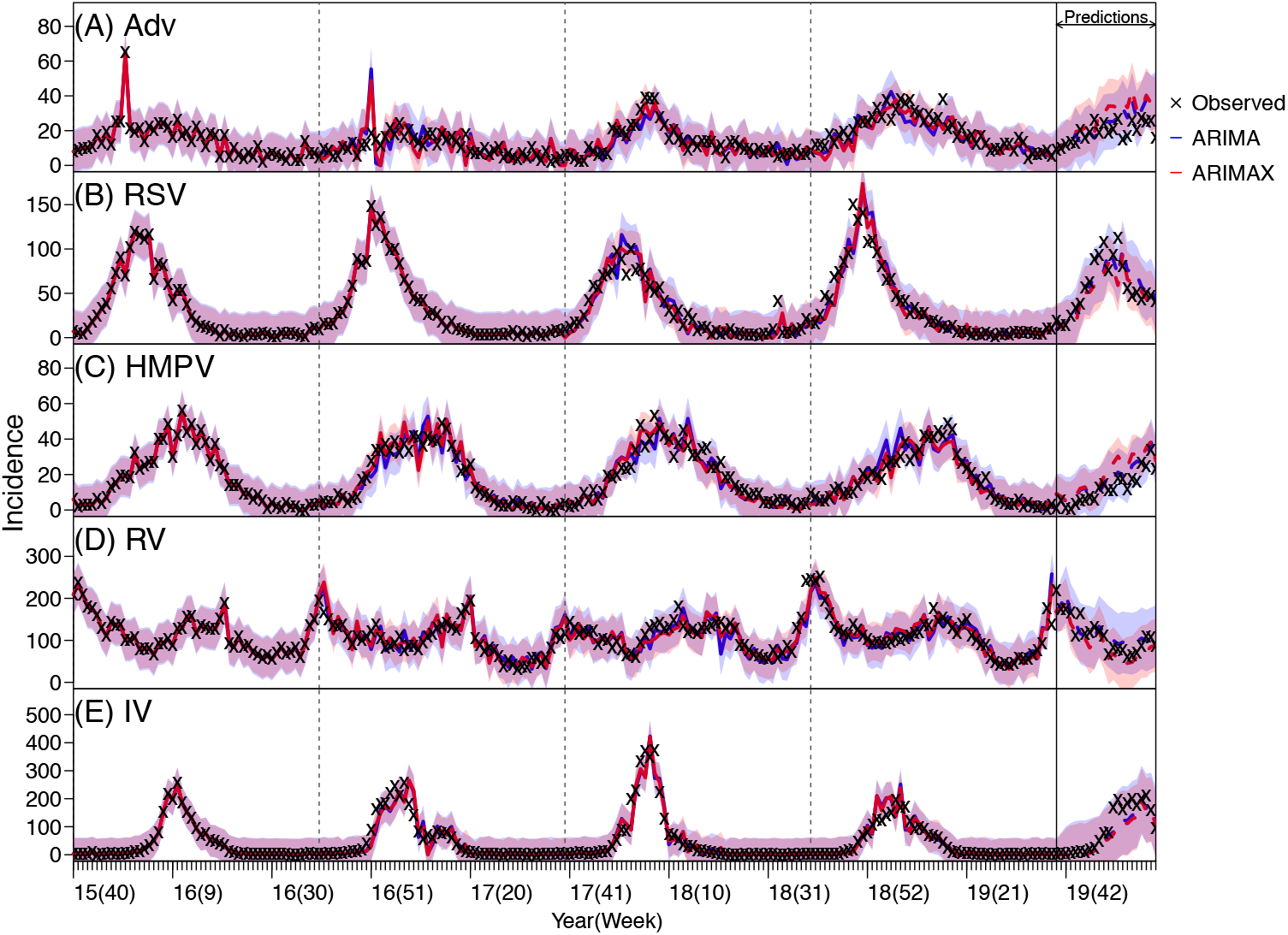
ARIMA and ARIMAX model validation for adenovirus (Adv; A), respiratory syncytial virus (RSV; B), human metapneumovirus (HMPV; C), rhinovirus (RV; D), and influenza virus (IV; E). ARIMA and ARIMAX models for each virus were first trained using incidence data from Week 40 of 2015 to Week 39 of 2019 and then used to predict incidence from Week 40 of 2019 to Week 9 of 2020. Crosses (‘x’) show scaled weekly incidence. Blue lines (mean) and shaded areas (95% confidence intervals) show model fit (solid lines) and prediction (dashed lines) using the ARIMA models; red lines (mean) and shaded areas (95% confidence intervals) show model fit (solid lines) and prediction (dashed lines) using the ARIMAX models.

**Fig S3.**
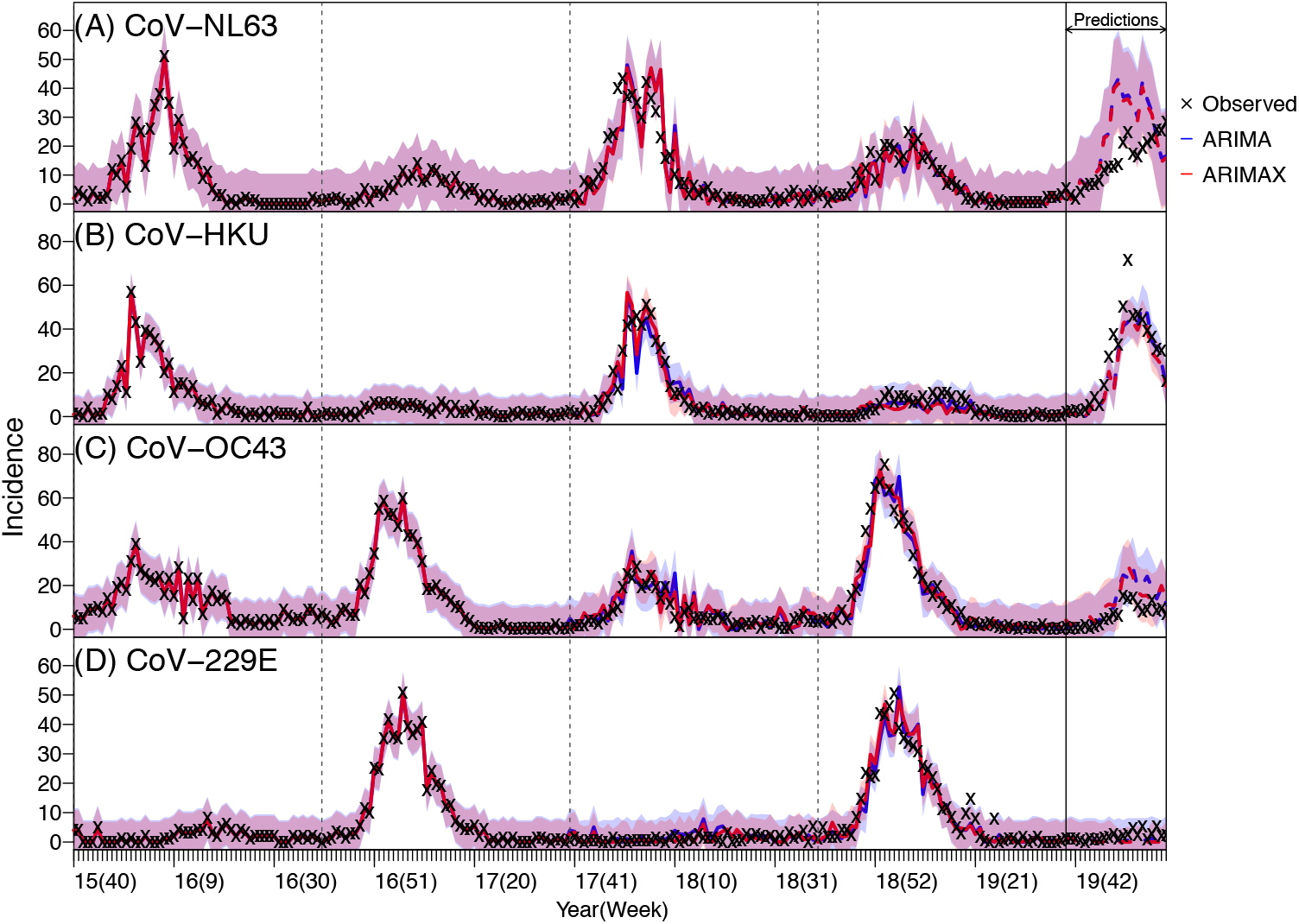
ARIMA and ARIMAX model validation for human endemic coronaviruses: CoV-NL63 (A), CoV-HKU (B), CoV-OC43 (C), and CoV-229E (D). ARIMA and ARIMAX models for each virus were first trained using incidence data from Week 40 of 2015 to Week 39 of 2019 and then used to predict incidence from Week 40 of 2019 to Week 9 of 2020. Crosses (‘x’) show scaled weekly incidence. Blue lines (mean) and shaded areas (95% confidence intervals) show model fit (solid lines) and prediction (dashed lines) using the ARIMA models; red lines (mean) and shaded areas (95% confidence intervals) show model fit (solid lines) and prediction (dashed lines) using the ARIMAX models.

**Fig S4.**
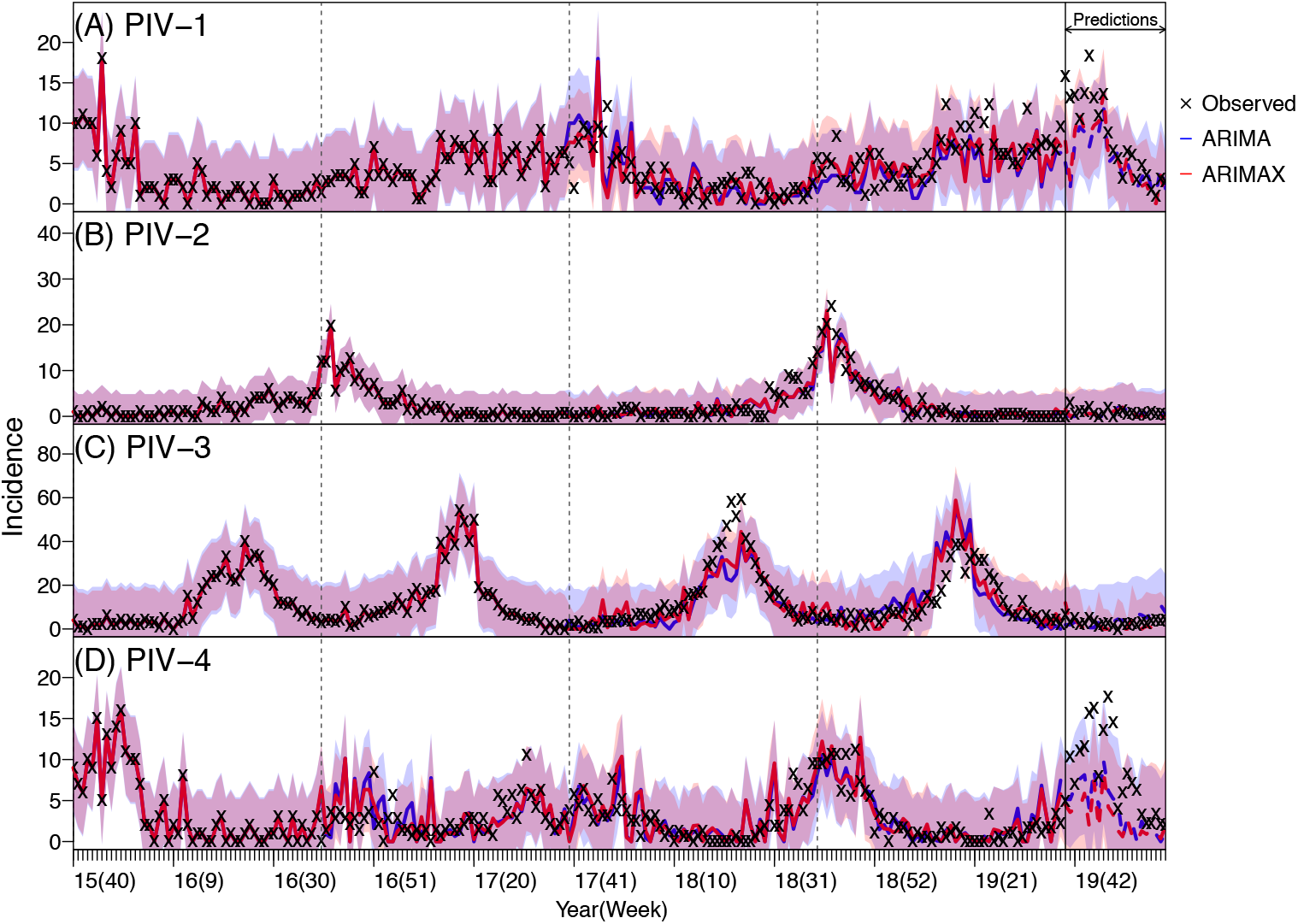
ARIMA and ARIMAX model validation for parainfluenza viruses: PIV-1 (A), PIV-2 (B), PIV-3 (C), and PIV-4 (D). ARIMA and ARIMAX models for each virus were first trained using incidence data from Week 40 of 2015 to Week 39 of 2019 and then used to predict incidence from Week 40 of 2019 to Week 9 of 2020. Crosses (‘x’) show scaled weekly incidence. Blue lines (mean) and shaded areas (95% confidence intervals) show model fit (solid lines) and prediction (dashed lines) using the ARIMA models; red lines (mean) and shaded areas (95% confidence intervals) show model fit (solid lines) and prediction (dashed lines) using the ARIMAX models.

